# Childhood Maltreatment and Risk for Illicit Substance Use: Evidence for Mid-Adolescence as a Sensitive Exposure Period

**DOI:** 10.1101/2025.10.10.25337769

**Authors:** Alaptagin Khan, Martin H. Teicher

## Abstract

**Background:** Childhood maltreatment is a well-established risk factor for substance misuse. However, it remains unclear whether risk for specific illicit substances is driven primarily by the cumulative burden of adverse experiences or by exposures during specific sensitive developmental periods.

**Methods:** We administered the Maltreatment and Abuse Chronology of Exposure (MACE) scale to 2,013 medically healthy young adults (693M/1320F, ages 18–25) to assess severity of exposure to ten maltreatment types across each year of childhood. Lifetime non-medical use of opioids, stimulants, cocaine, sedatives, hallucinogens, and polysubstance involvement was determined using the Alcohol, Smoking, and Substance Involvement Screening Test (ASSIST). We employed an integrative analytical approach, combining machine learning (random forests) with modern causal inference (Targeted Maximum Likelihood Estimation) and survival modeling.

**Findings:** Although maltreatment multiplicity (number of types) was associated with substance use in initial models, it was not a significant predictor in analyses that included specific type-time exposures. Machine learning and causal inference instead identified peer emotional abuse at age 15 in males and childhood sexual abuse at age 15 in females as potent risk factors for multiple outcomes, including opioid, stimulant, and polysubstance use. These specific exposures increased absolute risk by 20–35% (Risk Ratios: 2- to 4-fold), with associations robust to unmeasured confounding (E-values: 4–10). The apparent population-level dose-response relationship for multiplicity is therefore likely explained by the higher concentration of these critical type-time exposures among individuals with high maltreatment multiplicity.

**Interpretation:** Our findings challenge the cumulative burden model, providing robust evidence for mid-adolescence as a sensitive period during which specific maltreatment exposures, peer victimization in males and sexual abuse in females, causally shape trajectories of illicit substance misuse. Prevention strategies targeting these adversities and clinical screening during this developmental window may be crucial for reducing the population burden of substance use disorders.

**Funding:** National Institute on Drug Abuse USA (RO1 DA-017846), National Institute of Mental Health USA (RO1 MH-091391), the ANS Foundation, the Trauma Research Foundation, and Bessel van der Kolk.

## Introduction

Childhood adversity is a critical risk factor for substance use disorders (SUDs)^1^, though risk may vary based on the type of adversity. Kendler et al.,^2^, reported that non-genital childhood sexual abuse (CSA) was associated with a 2.93-fold increase in risk for drug dependence and that CSA involving intercourse was associated with a 5.70-fold increase.

Similarly, a comprehensive meta-analysis of CSA reported a 1.9 to 3.5-fold increase in drug use disorders^3^. In contrast, a detailed meta-analysis of non-sexual forms of childhood maltreatment reported 1.92-, 1.41-, and 1.36-fold increased risk for drug use with physical abuse, emotional abuse, and neglect, respectively^4^. These associations justify a rigorous effort to understand more precisely how maltreatment acts to increase risk.

While we know that maltreatment increases risk, we do not know whether the critical determinant is the cumulative burden of exposure to multiple types of maltreatment^1^, exposure to specific types of abuse during sensitive periods^5^, overall severity of abuse^5,6^, or recency of exposure^7^. The Adverse Childhood Experiences (ACEs) study provides evidence for a dose-dependent relationship, with exposure to more ACEs resulting in increased risk^1^. While this relationship is clear, the interpretation that the determining factor is “*cumulative exposure to traumatic stress during childhood”*^8^ is not. The alternative hypothesis is that risk is better attributed to exposure to specific types of abuse during narrow sensitive periods. The apparent increase in risk with increasing exposure may be a byproduct of the fact that exposure to more types of maltreatment increases the likelihood of experiencing a critical type of abuse at a critical age^5^.

A sensitive period hypothesis has considerable translational appeal as brain regions become sensitive to environmental factors during specific stages of development when inhibitory and excitatory neurotransmitter systems come into balance, and these periods of plasticity are then attenuated by molecular breaks^9^.

Several studies have provided support for potential sensitive period effects of maltreatment on multiple brain regions (e.g., hippocampus^10^, amygdala^11^, prefrontal cortex^12^ , and corpus callosum^13^) and clinical outcomes (e.g., depression and suicidal ideation^5^, positive and negative psychotic symptoms^14^, cognitive deficits^15^). In these instances, exposure to a specific type of maltreatment during a brief period was a significantly more important risk factor than multiplicity or severity of maltreatment across childhood. However, there were other outcomes (e.g., dissociation and risk for PTSD) where severity or multiplicity were the most important risk factors^5,16^. Hence, this is an empirical question that needs to be determined for each outcome.

We also found that sensitive periods can be sex-specific. For example, we reported that the most important risk factor for reduced hippocampal volume was neglect during the first seven years of life in males and abuse at ages 10, 11, 15, and 16 in females^10^. In other instances, such as risk for major depression, we found that males and females were vulnerable during the same ages but to different types of maltreatment^5^. More recently, we reported that brain regions can have multiple sensitive periods, which may be associated with opposite effects on brain function. For instance, exposure to physical abuse between 3 and 6 years of age was associated in early adulthood with a blunted amygdala response to threat, whereas peer emotional bullying at 13 and 15 years was associated with an augmented response^11^.

Whether risk is best explained by sensitive periods, cumulative burden, or overall severity of exposure is of fundamental concern. From a cumulative burden perspective, reducing exposure to any adversity would diminish risk to a slight degree. In contrast, a sensitive period perspective has the potential to identify types of adversity at specific ages, which, if prevented, might markedly reduce risk. Therefore, this study aimed to ascertain the importance of type and timing of maltreatment versus cumulative exposure in delineating risk for illicit substance use in emerging adults. We predicted that the data would be more supportive of a sensitive period than a cumulative burden model. In addition, we assessed using causal inference analysis whether there was a potential causal relationship between exposure to specific adverse experiences and risk for drug use, rigorously evaluated these findings through an array of sensitivity analyses, and used survival analyses to provide information on the developmental time course of escalating use.

## Method

This Project was reviewed and approved by the Mass General Brigham IRB, Assurance # FWA00002744. All participants gave written informed consent before participation.

### Participants

Subjects were recruited as part of an online screening process for potential inclusion into two NIH-funded studies with neuroimaging components. Participants were required to be medically healthy, currently unmedicated, right-handed, and between 18-25 years of age. Recruitment was from metropolitan and suburban areas around Boston, Massachusetts through advertisement, followed by a phone screening. Participants who passed the phone screen were given a password to a HIPAA-compliant online enrollment system, which collected detailed information on their life experiences, medical and psychiatric history, developmental history, demographics, psychiatric symptomatology, substance use, and MACE scale. Of the 2,013 participants, providing complete online maltreatment and substance use data, 656 attended in-office visits at McLean Hospital that included clinical diagnostic interviews, neurocognitive assessments, and neuroimaging (N = 342). Online data were collected between 11/8/2010 and 12/9/2016; well in advance of the COVID-19 pandemic.

### Demographics

Self-report data were collected on race, ethnicity, sex at birth, education, parental education, family income, and perceived financial sufficiency during childhood (1=much less than enough money to meet our needs, to 5=much more than enough money to meet our needs). We included perceived financial sufficiency in addition to family income, as participants were often uncertain of their parents’ income during childhood. In previous studies, perceived financial sufficiency explained a greater share of the variance in symptom ratings than family income and percent of poverty level.

### Maltreatment and Abuse Chronology of Exposure Scale (MACE)

The severity of exposure to maltreatment across each year of childhood was assessed retrospectively using the 75-item MACE-X^17^. The ten categories of maltreatment assessed included physical and sexual abuse, parental verbal and non-verbal emotional abuse, physical and emotional neglect, witnessing interparental violence, witnessing violence to siblings, and peer physical and emotional bullying. By meeting criteria for the dichotomous Rasch model, each MACE category provided a ‘fundamental measure’ of exposure with at least interval scaling properties. MACE scores have high overall test-retest reliability (r=0.91^17^, ICC ≥ 0.94^18^), good-to-excellent reliability at specific ages, and do not show significant negative attribution bias^17^. The MACE has good convergent validity (r= .738) with the childhood trauma questionnaire CTQ and (r=.698) and ACEs scale, but accounted for 2.00- and 2.07-fold more of the variance in symptom ratings using variance decomposition analyses. The in-office sample was also administered the Traumatic Antecedents Interview^19^

### Illicit Substance Use

Substance use was assessed using an abbreviated version of the World Health Organization Alcohol, Smoking, and Substance Involvement Screening Test (ASSIST)^20^, which focused on lifetime history of use and use during the past three months. This is a frequently used measure in epidemiology studies and is available free of charge.

### Sensitive Periods Analyses

Identifying sensitive periods is complex given the collinearity in maltreatment ratings. We found in a series of Monte-Carlo simulations that a specific machine learning strategy using random forest (RF) regression^21^ with conditional inference trees^22^ was particularly successful in identifying the underlying predictor variables in simulated data sets.

Briefly, a forest of 1000 decision trees was created for each outcome from different subsets of the data and constrained in the number of predictors considered at each decision point. Pooling the results from all the decision trees provides a ‘wisdom of the crowd’ strategy that is resistant to collinearity, can model interactions, does not assume a linear exposure-response relationship, and typically provides better predictions than conventional approaches.

The dataset was randomly split into training (63.3% of the participants) and test (36.7%) sets. Each risk factor’s variable importance (VI) was assessed by sequentially permuting the risk factor and refitting the model to determine the degree to which the loss of information due to permutation worsened the fit^21,22^. VI was defined as the increase in mean square error (MSE) following permutation and averaged over 50 random splits of the data. Permuting important predictors increased MSE to a large degree, whereas permuting unimportant predictors had little impact. The significance of the VI measures was determined by Z-tests with Bonferroni correction by comparing actual VI measures to VI measures derived from reshuffled data repeated 1000 times. One limitation of RF analyses is that they indicate the importance of the risk factor, but the relationship between the risk factor and the outcome is unclear. To assess this relationship, we used the saved random forest to determine the dose-response relationship between changes in the risk factor of interest and the predicted outcome, holding all the other risk factors at their median level, which is facilitated using RF regression rather than RF classification.

Type/time risk factors experienced by at least 2% of participants, along with additional covariates of age, parental education, financial sufficiency, race, maternal and paternal histories of drug abuse, and alcohol abuse, were included in the analyses. Penalized regression analyses were used to verify that significant risk factors identified by random forest were included in the limited subset of variables selected by the RF.

### Statistical Criteria for Sensitive Exposure Periods

We previously proposed^5^ that a sensitive exposure period can be said to exist if exposure to maltreatment during a specific developmental stage was a more important risk factor than overall measures of childhood exposure, as indexed by the severity, duration, and multiplicity of exposure experienced throughout childhood.

### Causal Analysis of Maltreatment and Substance Misuse

To estimate the causal effects of childhood maltreatment type and timing on subsequent substance misuse, we employed a counterfactual-based analytic framework that integrates multiple imputation (MI) for missing data with Targeted Maximum Likelihood Estimation (TMLE) for robust effect estimation. This approach was selected for its efficiency, reduced sensitivity to model misspecification, and ability to accommodate realistic missing data scenarios.

Missingness was concentrated in four parental history variables (maternal and paternal alcohol and drug misuse). Data were multiply imputed under the missing-at-random (MAR) assumption using predictive mean matching (mice package, 20 imputations). Results were pooled across imputations using Rubin’s rules. To address the possibility of missing-not-at-random (MNAR) bias (e.g., underreporting of parental substance problems), we conducted sensitivity analyses by inflating the odds of parental history in the imputed datasets by 1.5-fold, 2-fold, and 3-fold, thereby testing the robustness of estimates to varying degrees of underreporting.

Within each imputed dataset, we applied Targeted Maximum Likelihood Estimation (TMLE) using the *tmle* R package. TMLE is a doubly robust, semi-parametric method that combines machine learning–based estimation of the exposure and outcome processes with a targeting step that reduces bias while controlling variance. In the first stage, TMLE fits a model for the probability of exposure (propensity score) and a model for the conditional outcome regression, incorporating covariates such as parental education, household financial status, parental substance history, race/ethnicity, prior exposure to the same adversity type (lagged by three years to distinguish prior from concurrent exposure), and number of different types of maltreatment experienced (excluding the specific type being assessed). These initial models were estimated using Super Learner, an ensemble approach that combines multiple candidate algorithms through cross-validation to optimize predictive accuracy. In the second stage, TMLE applies a fluctuation, or “targeting,” step that updates the initial outcome regression using information from the estimated propensity score. This adjustment aligns the estimator with the parameter of interest (e.g., risk difference or risk ratio), ensuring that it solves the efficient influence function equation for the target causal parameter. As a result, TMLE produces doubly robust estimates, consistent if either the exposure or outcome model is correctly specified, and attains the semiparametric efficiency bound when both are correctly specified. This framework leverages flexible machine learning ensembles to improve robustness to model misspecification while maintaining valid statistical inference. To assess the sensitivity of the effects, analyses were repeated under three conditions: (1) complete-case data only, (2) MI under MAR, and (3) MNAR-adjusted imputations. Covariate balance between exposed and unexposed groups was assessed using standardized mean differences (tableone, survey packages). No covariate exhibited a significant residual imbalance after adjustment.

Primary causal effect measures were the risk difference (RD) and risk ratio (RR). RD quantifies the absolute excess risk attributable to maltreatment and is most relevant for public health planning, while RR highlights proportional increases in risk across populations with different baselines. To assess the robustness of findings to potential unmeasured confounding, we also calculated E-values for RD and RR estimates, as well as for the confidence limits of RR. The E-value represents the minimum strength of association that an unmeasured confounder would need to have with both the exposure and the outcome, beyond the measured covariates, to fully explain away the observed association. In general, E-values greater than 2 suggest moderate robustness, while values above 4–5 indicate that only a very strong, and often implausible, unmeasured confounder could account for the observed effect. E-values were also calculated for the **l**ower limit of the confidence interval (closest to the null). If the CI-limit E-value is still large (e.g., >2), then even under the weakest effect consistent with the data, the result remains robust to unmeasured confounding.

### Survival Analyses

To complement the TMLE analysis and specifically examine the causal effect of maltreatment on the *timing* of substance misuse onset, we conducted survival analyses using a weighted Cox proportional hazards model. This approach allowed for the estimation of hazard ratios while appropriately accounting for right-censored data. To adjust for confounding variables and maintain a consistent causal inference framework, we employed inverse probability weighting (IPW). First, we estimated propensity scores, the probability of exposure to a specific maltreatment, for each participant using a logistic regression model that included the full set of covariates in females, and the full set minus the family history covariate in males. We chose this approach as none of the family history variables were significant in males, and several of these variables were missing. To ensure model stability, propensity scores were clipped to prevent values of exactly 0 or 1. From these scores, we calculated stabilized inverse probability weights, which were then truncated at the 99th percentile to minimize the influence of extreme outlier weights. After confirming successful covariate balance in the weighted sample using standardized mean differences, we fitted a weighted Cox proportional hazards model using the svycoxph function in the survey package. Finally, weighted Kaplan-Meier curves were generated using svykm and plotted with ggplot2 to visually represent the difference in the probability of remaining free of substance misuse over time between the exposed and unexposed groups.

### Role of the funding source

The funding sources (NIMH, NIDA, ANS foundation) had no role in the study design, data collection, analysis, and interpretation of data, writing of the report, or in the decision to submit the paper for publication.

## Results

### Sample

Of the initial cohort (N=2092), 2013 participants (693M/1320F; 96.2%) provided complete MACE and substance use data (mean age = 22.3 ± 2.2 years). Demographics are shown in Table 1.

**TABLE 1.**
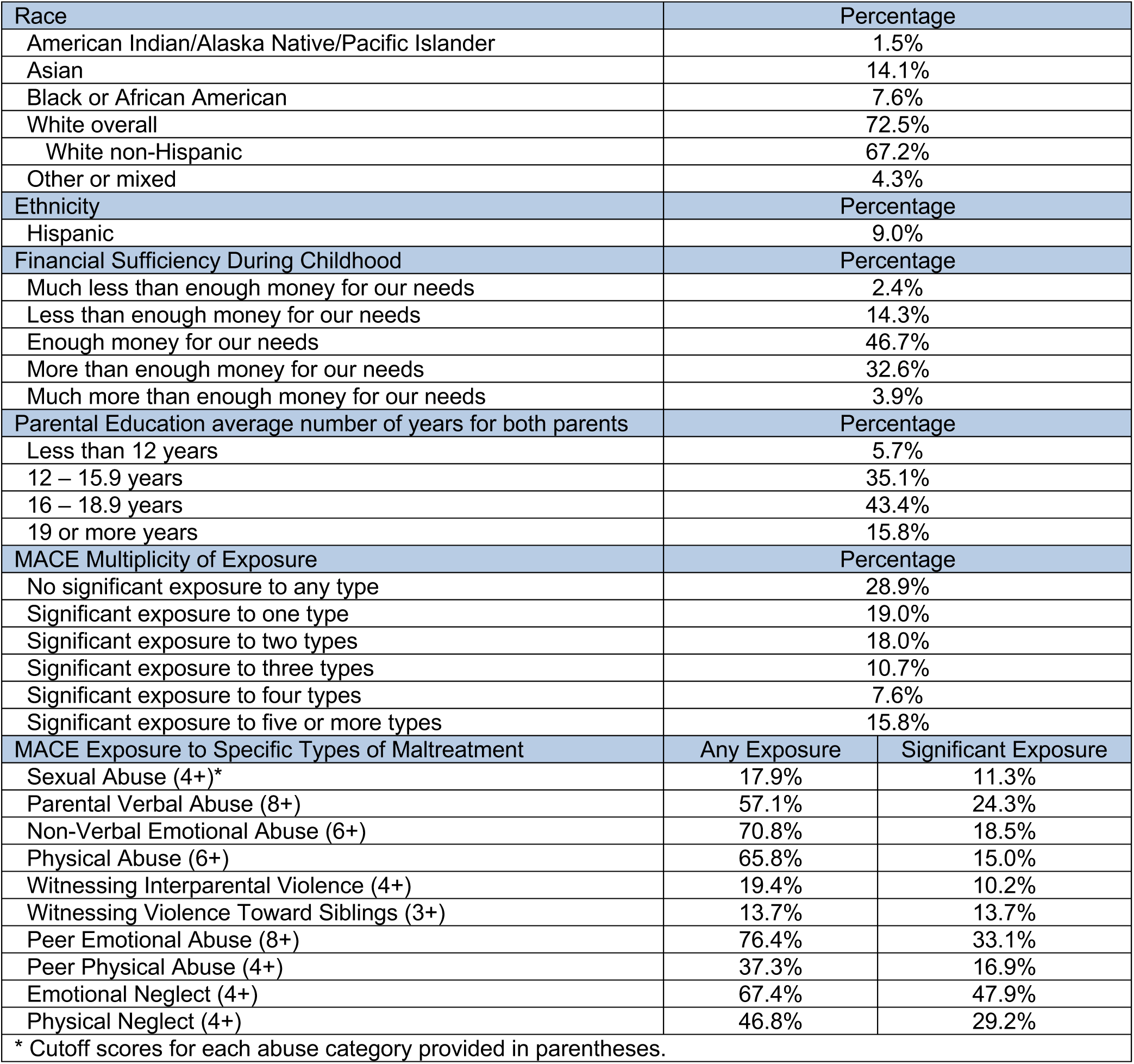
Demographics and maltreatment history N = 2013.

| Race | Percentage |  |
| --- | --- | --- |
| American Indian/Alaska Native/Pacific Islander | 1.5% |  |
| Asian | 14.1% |  |
| Black or African American | 7.6% |  |
| White overall | 72.5% |  |
| White non-Hispanic | 67.2% |  |
| Other or mixed | 4.3% |  |
| Ethnicity | Percentage |  |
| Hispanic | 9.0% |  |
| Financial Sufficiency During Childhood | Percentage |  |
| Much less than enough money for our needs | 2.4% |  |
| Less than enough money for our needs | 14.3% |  |
| Enough money for our needs | 46.7% |  |
| More than enough money for our needs | 32.6% |  |
| Much more than enough money for our needs | 3.9% |  |
| Parental Education average number of years for both parents | Percentage |  |
| Less than 12 years | 5.7% |  |
| 12 – 15.9 years | 35.1% |  |
| 16 – 18.9 years | 43.4% |  |
| 19 or more years | 15.8% |  |
| MACE Multiplicity of Exposure | Percentage |  |
| No significant exposure to any type | 28.9% |  |
| Significant exposure to one type | 19.0% |  |
| Significant exposure to two types | 18.0% |  |
| Significant exposure to three types | 10.7% |  |
| Significant exposure to four types | 7.6% |  |
| Significant exposure to five or more types | 15.8% |  |
| MACE Exposure to Specific Types of Maltreatment | Any Exposure | Significant Exposure |
| Sexual Abuse (4+)* | 17.9% | 11.3% |
| Parental Verbal Abuse (8+) | 57.1% | 24.3% |
| Non-Verbal Emotional Abuse (6+) | 70.8% | 18.5% |
| Physical Abuse (6+) | 65.8% | 15.0% |
| Witnessing Interparental Violence (4+) | 19.4% | 10.2% |
| Witnessing Violence Toward Siblings (3+) | 13.7% | 13.7% |
| Peer Emotional Abuse (8+) | 76.4% | 33.1% |
| Peer Physical Abuse (4+) | 37.3% | 16.9% |
| Emotional Neglect (4+) | 67.4% | 47.9% |
| Physical Neglect (4+) | 46.8% | 29.2% |
| * Cutoff scores for each abuse category provided in parentheses. |  |  |

### Multiplicity and Dose-Response

Consistent with prior ACEs findings, risk of illicit substance use increased with exposure to a greater number of maltreatment types (Supplemental Figure 1, Table S1). For example, adjusted odds of polysubstance use were 1.81-fold (95% CI, 1.18–2.78), 2.17-fold (95% CI, 1.33–3.54), and 2.86-fold (95% CI, 1.93–4.26) higher in participants reporting 3, 4, or ≥5 types of maltreatment, respectively, compared to those reporting none (χ²(5)=33.2, p<.0001). These findings are very much in line with prior reported studies and support the comparison of the cumulative burden model versus a sensitive period model in this sample.

### Random Forest Identification of Type/Time Predictors

As indicated in Table 2 and Figure S2, results from the Random Forest regression in males revealed that the risk for illicit substance misuse was not predicted by cumulative measures of adversity, such as the number of different maltreatment types experienced (MACE Multiplicity). Instead, the analysis identified highly specific risk profiles where the type and developmental timing of the exposure were the most salient predictors.

**TABLE 2.** Significance of type/time risk factors and measures of overall exposure associated with specific classes of illicit drug use in males and females based on random forest regression.

| MALES |  |  |  |  | FEMALES |  |  |  |
| --- | --- | --- | --- | --- | --- | --- | --- | --- |
| Drug Category | Risk Factors | Age | VI | Adjusted p-value | Risk Factors | Age | VI | Adjusted p-value |
| Polysubstance use | Peer Emotional | 15 | 0.48 | 0.03 | Sexual Abuse | 15 | 0.96 | <10<sup>-16</sup> |
|  | Financial Sufficiency |  | 2.83 | <10<sup>-16</sup> | Sexual Abuse | 17 | 0.33 | 0.006 |
|  | MACE Multiplicity |  | 0.07 | NS | Mother's Alcohol |  | 0.35 | 0.03 |
|  | MACE Severity |  | 0.06 | NS | MACE Multiplicity |  | 0.11 | NS |
|  |  |  |  |  | MACE Severity |  | 0.24 | NS |
| Opiates | Peer Emotional | 15 | 1.42 | <10<sup>-16</sup> | Peer Emotional | 15 | 1.02 | <10<sup>-9</sup> |
|  | Peer Emotional | 16 | 0.57 | 0.00063 | Peer Emotional | 18 | 0.66 | <10<sup>-5</sup> |
|  | Peer Emotional | 18 | 0.60 | 0.00077 | Physical Neglect | 12 | 0.50 | 0.013 |
|  | Peer Physical | 16 | 0.64 | <10<sup>-8</sup> | Parental Verbal | 2 | 0.11 | 0.0026 |
|  | Age |  | 0.79 | 0.018 | Parental Verbal | 3 | 0.31 | <10<sup>-10</sup> |
|  | MACE Multiplicity |  | 0.07 | NS | Witness IPV | 9 | 0.23 | 0.00024 |
|  | MACE Severity |  | 0.03 | NS | MACE Multiplicity |  | 0.04 | NS |
|  |  |  |  |  | MACE Severity |  | 0.27 | NS |
| Stimulants | Peer Emotional | 15 | 0.56 | 0.014 | Sexual Abuse | 15 | 1.16 | <10<sup>-16</sup> |
|  | Financial Sufficiency |  | 1.96 | <10<sup>-15</sup> | Sexual Abuse | 18 | 0.57 | <10<sup>-14</sup> |
|  | MACE Multiplicity |  | 0.05 | NS | Witness Viol Sib | 8 | 0.23 | <10<sup>-5</sup> |
|  | MACE Severity |  | 0.035 | NS | Peer Emotional | 11 | 0.57 | 0.0091 |
|  |  |  |  |  | Mother's Alcohol |  | 0.39 | <10<sup>-4</sup> |
|  |  |  |  |  | MACE Multiplicity |  | 0.22 | NS |
|  |  |  |  |  | MACE Severity |  | 0.35 | NS |
| Sedatives | Peer Emotional | 15 | 0.66 | 0.01 | Emotional Neglect | 9 | 0.55 | <10<sup>-7</sup> |
|  | Peer Emotional | 17 | 0.76 | <10<sup>-7</sup> | Sexual Abuse | 15 | 0.40 | <10<sup>-9</sup> |
|  | MACE Multiplicity |  | 0.035 | NS | Sexual Abuse | 18 | 0.42 | 0.0017 |
|  | MACE Severity |  | 0.042 | NS | Parental Verbal | 3 | 0.18 | 0.0089 |
|  |  |  |  |  | Peer Physical | 15 | 0.47 | <10<sup>-16</sup> |
|  |  |  |  |  | Peer Physical | 16 | 0.24 | <10<sup>-4</sup> |
|  |  |  |  |  | Peer Physical | 18 | 0.21 | 0.017 |
|  |  |  |  |  | Mother's Alcohol |  | 0.76 | <10<sup>-16</sup> |
|  |  |  |  |  | MACE Multiplicity |  | 0.97 | <10<sup>-6</sup> |
|  |  |  |  |  | MACE Severity |  | 0.60 | NS |
| Hallucinogens | Witness Viol Sib | 17 | 0.18 | <10<sup>-16</sup> | Sexual Abuse | 15 | 0.89 | <10<sup>-16</sup> |
|  | Sexual Abuse | 16 | 0.15 | <10<sup>-13</sup> | Sexual Abuse | 17 | 0.34 | 0.04 |
|  | Sexual Abuse | 18 | 0.24 | <10<sup>-16</sup> | Father's Alcohol |  | 0.67 | <10<sup>-5</sup> |
|  | Financial Sufficiency |  | 3.18 | <10<sup>-16</sup> | MACE Multiplicity |  | 0.04 | NS |
|  | MACE Multiplicity |  | 0.06 | NS | MACE Severity |  | 0.1 | NS |
|  | MACE Severity |  | 0.03 | NS |  |  |  |  |
|  | Race |  | 0.71 | 0.007 |  |  |  |  |
| Cocaine | Sexual Abuse | 16 | 0.10 | 0.011 | Sexual Abuse | 15 | 1.12 | <10<sup>-16</sup> |
|  | Financial Sufficiency |  | 0.96 | <10<sup>-5</sup> | Peer Emotional | 17 | 0.50 | 0.0050 |
|  | MACE Multiplicity |  | 0.09 | NS | Peer Physical | 15 | 0.24 | <10<sup>-4</sup> |
|  | MACE Severity |  | 0.10 | NS | Mother's Alcohol |  | 0.57 | <10<sup>-6</sup> |
|  |  |  |  |  | MACE Multiplicity |  | 0.08 | NS |
|  |  |  |  |  | MACE Severity |  | 0.12 | NS |

IPV – Interparental Violence, NS – not significant, VI – Variable Importance, Viol – Violence

Specifically, exposure to peer emotional abuse during mid-to-late adolescence, particularly age 15, emerged as a powerful risk factor for polysubstance, opiate, stimulant, and sedative use. In contrast, the strongest risk factors for hallucinogen and cocaine use were exposure to sexual abuse during later adolescence (ages 16 and 18). These findings suggest that for males, the neurodevelopmental impact of specific forms of maltreatment during sensitive periods of adolescence is a more critical determinant of substance misuse risk than the overall cumulative burden of adversity. Financial sufficiency also emerged as a significant risk factor for polysubstance, stimulant, hallucinogen, and cocaine use in males, but not females, with increased risk associated with high levels of perceived financial sufficiency (Figure S3).

Similar to the findings in males, the analysis in females indicated that specific maltreatment experiences at sensitive developmental periods were more salient risk factors for substance misuse than the overall MACE Multiplicity score, with the notable exception of sedative use (Table, Fig S4). However, the specific risk profiles for females were distinct and often more complex. Sexual abuse during mid-adolescence (age 15) was a more pervasive and powerful predictor for females than for males, emerging as a top risk factor across polysubstance use, stimulants, sedatives, hallucinogens, and cocaine. Furthermore, while the risk for males was largely confined to adolescent peer and sexual abuse, the risk profile for females was broader, encompassing earlier life adversities such as parental verbal abuse in early childhood (ages 2-3) and witnessing violence in middle childhood, particularly for opiate and sedative misuse. Parental substance use also appeared as a significant predictor for females across multiple drug categories, a factor that was not identified for males.

### Causal Inference Analyses

Targeted Maximum Likelihood Estimation (TMLE) incorporating Super Learner provided robust evidence for the causal impact of specific maltreatment exposures during sensitive developmental windows on lifetime substance misuse, as presented in Table 3. Results were consistent across complete-case and MNAR sensitivity analyses (inflating parental substance history by 1.5–3×), and moderate-to-large E-values (≈4–10) indicated that only strong unmeasured confounding could plausibly explain away the observed effects.

**TABLE 3.**
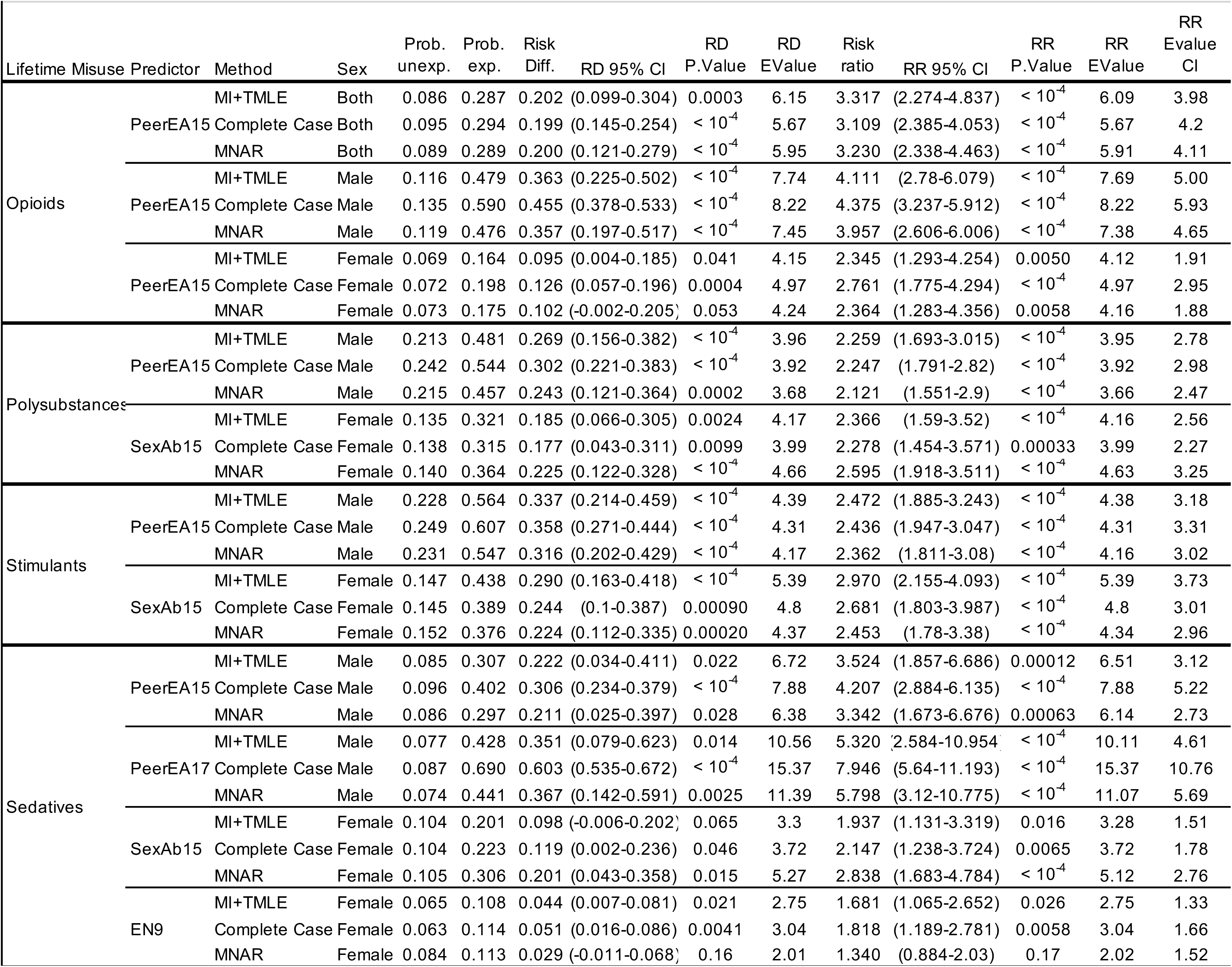
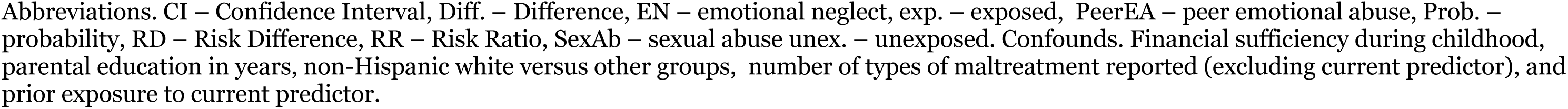
Causal Inference from Targeted Maximum Likelihood Estimation after Multiple Imputation with Sensitivity Tests.

### Opiate Misuse

Exposure to peer verbal abuse at age 15 was a potent causal risk factor for subsequent opiate misuse. In the combined sample, this exposure was associated with an absolute increase in risk of 20.2% (Risk Difference [RD] = 0.20, 95% CI: 0.10 to 0.30, p < .001) and more than tripled the relative risk (Risk Ratio [RR] = 3.32, 95% CI: 2.27 to 4.84, p < .001). The effect was particularly strong in males, who experienced a 36.3% absolute increase in risk (RD = 0.36, 95% CI: 0.22 to 0.50) and a four-fold increase in relative risk (RR = 4.11, 95% CI: 2.78 to 6.08). The effect was also significant but more modest in females (RD = 9.5%, RR = 2.34).

These findings were highly robust to unmeasured confounding, with E-values for the risk ratio point estimates of 7.69 for males and 4.12 for females.

### Polysubstance and Stimulant Misuse

Distinct maltreatment pathways were identified for polysubstance and stimulant misuse between males and females. For males, peer verbal abuse at age 15 was the key risk factor, increasing the absolute risk of polysubstance use by 26.9% (RR = 2.26) and stimulant use by 33.7% (RR = 2.47). For females, the primary causal factor was sexual abuse at age 15, which increased the absolute risk of polysubstance use by 18.5% (RR = 2.37) and stimulant use by 29.0% (RR = 2.97). All of these associations were highly statistically significant (p < .001) and robust to unmeasured confounding, with E-values for the risk ratios ranging from 3.95 to 5.40.

### Sedative Misuse

Risk for sedative misuse was associated with multiple distinct exposures. In males, peer verbal abuse at age 15 increased absolute risk by 22.2% (RR = 3.52), while exposure at age 17 conferred an even greater risk, increasing absolute risk by 35.1% (RR = 5.32). In females, risk was primarily linked to sexual abuse at age 15 (RD = 9.8%, RR = 1.94) and emotional neglect at age 9 (RD = 4.4%, RR = 1.68).

### Hallucinogen and Cocaine Misuse

Exposure to sexual abuse during adolescence was the primary causal risk factor for hallucinogen and cocaine misuse, though the specific timing differed by sex. For females, sexual abuse at age 15 significantly increased the risk for both hallucinogen misuse (RD = 18.4%, RR = 2.52) and cocaine misuse (RD = 17.0%, RR = 3.10). For males, the critical exposure period was later; sexual abuse at age 16 and 18, along with witnessing sibling abuse at age 17, were strong causal factors for hallucinogen misuse.

Notably, while sexual abuse at age 16 was identified by the Random Forest analysis as a predictor for cocaine misuse in males, the MI+TMLE analysis did not find a statistically significant causal effect for this specific pathway (RD = 33.5%, p = 0.247), suggesting the association may be explained by other measured covariates.

### Survival Analyses

IPW-adjusted Cox proportional hazards models confirmed that specific childhood maltreatment experiences significantly increased the hazard of initiating illicit substance use during adolescence and young adulthood. As illustrated by the weighted Kaplan-Meier survival curves (Figure 1), the timing of this increased risk showed distinct developmental patterns. For males, the survival curves for exposed and unexposed groups typically began to diverge around age 20 (or age 19 for hallucinogens), with this risk gap widening until approximately age 23. In females, the divergence in risk generally began between ages 20-22 and continued to increase throughout the observation period. This temporal pattern supports the directionality of the effect, demonstrating a latency of several years between the adolescent maltreatment exposure and the manifestation of substance use risk in early adulthood.

**FIGURE 1.**
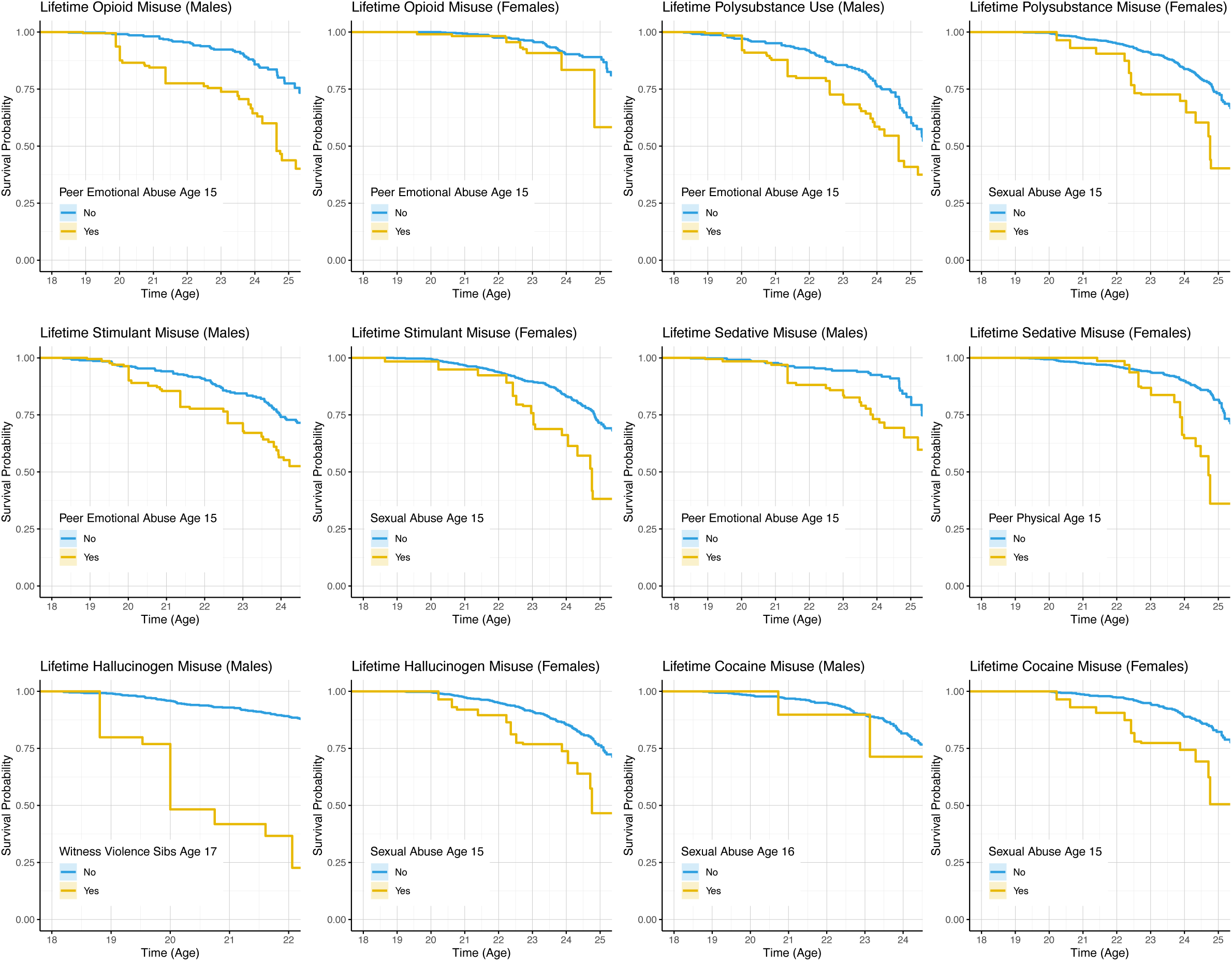
Inverse Probability Weighted Cox-Proportional Hazard Models showing differences in rates of survival (absence of non-medical use) across substance classes for males and females. Data were adjusted for parental education and perceived financial sufficiency across childhood, number of different types of maltreatment reported (excluding the type being analyzed), and prior exposure to the type of maltreatment being analyzed. Data for females were also adjusted for maternal and paternal history of drug and alcohol abuse.

The statistical results from the Cox models revealed strong and significant causal effects (Table S1). The majority of the hazard ratios ranged from 1.94 to 3.57, with the most extreme finding indicating a more than 12- fold increase in risk (HR = 12.12). Except for two comparisons that approached but did not reach statistical significance, opioid use following peer emotional abuse at age 15 in females (p = 0.083) and polysubstance use following the same exposure in males (p = 0.064), all other primary maltreatment pathways identified by the Random Forest analysis were confirmed to have a significant causal effect on the timing of substance use initiation.

### Validation of Self-Report

As indicated in Table 4, we found in a subsample with structured interviews (N=656), that self-reported substance use strongly predicted DSM-IV-TR substance use disorder diagnoses, with the highest odds for cocaine (aOR = 10.5) and opioid use (aOR = 8.4).

**TABLE 4.**
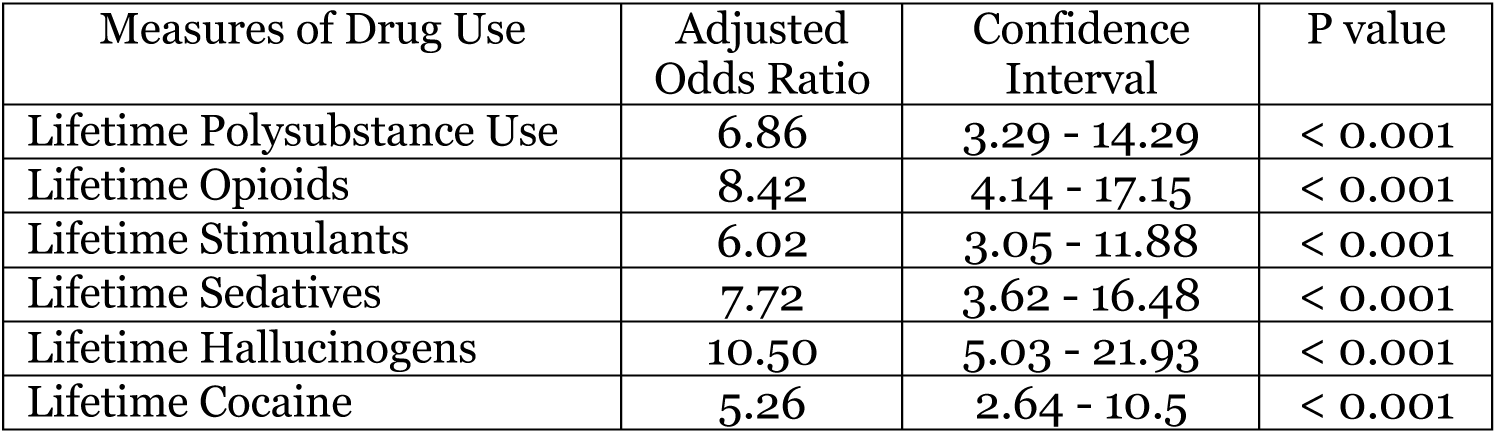
Relationship between self-reported drug use and adjusted odds of a clinical diagnosis of a substance use disorder on structured diagnostic interview. Data adjusted for age, sex, perceived financial sufficiency, race-ethnicity.

| Measures of Drug Use | Adjusted Odds Ratio | Confidence Interval | P value |
| --- | --- | --- | --- |
| Lifetime Polysubstance Use | 6.86 | 3.29 - 14.29 | < 0.001 |
| Lifetime Opioids | 8.42 | 4.14 - 17.15 | < 0.001 |
| Lifetime Stimulants | 6.02 | 3.05 - 11.88 | < 0.001 |
| Lifetime Sedatives | 7.72 | 3.62 - 16.48 | < 0.001 |
| Lifetime Hallucinogens | 10.50 | 5.03 - 21.93 | < 0.001 |
| Lifetime Cocaine | 5.26 | 2.64 - 10.5 | < 0.001 |

## Discussion

This study confirms that exposure to multiple types of childhood maltreatment is associated with an elevated risk of illicit substance use ^1,23–26^. However, by integrating machine learning, causal inference, and survival modeling, our findings challenge the cumulative burden model as the dominant explanatory framework. Instead, we demonstrate that specific type–time exposures during mid-adolescence exert the strongest causal influence on subsequent substance misuse, whereas global measures of multiplicity and severity were not significantly predictive once these sensitive periods were accounted for.

Our analytical approach provides robust, multi-faceted evidence for this conclusion. The use of Targeted Maximum Likelihood Estimation (TMLE) with Super Learner, a doubly robust framework, minimized model misspecification and confirmed that peer emotional abuse at age 15 in males and childhood sexual abuse at age 15 in females were potent and specific predictors. The associated risk ratios, typically in the two- to four-fold range, were underscored by E-values between 4 and 10, indicating that only strong, unmeasured confounding could plausibly account for these associations. Complementing this, IPW-weighted Cox proportional hazards models revealed that these adolescent exposures not only increased overall risk but also accelerated the timing of substance use onset, with hazard ratios in the two- to three-fold range. The integration of these distinct analytic frameworks, each robust to different assumptions, increases confidence in the validity of the findings and provides strong evidence that maltreatment during mid-adolescence plays a direct causal role in shaping trajectories of substance misuse.

These findings have important theoretical implications, aligning with a sensitive period framework in which neural systems undergoing developmental remodeling confer heightened vulnerability to environmental insults. Mid-adolescence appears to be a critical period for exposures that may disrupt the maturation of emotion regulation and reward circuits. The apparent dose–response relationship between maltreatment multiplicity and risk is likely driven by the increased probability that individuals with high multiplicity also experienced the most etiologically consequential exposures. For example, the percentage of females reporting any degree of CSA at age 15 increased monotonically from 0% to 21% with increasing MACE MULTIPLICITY scores, and the percentage of males reporting clinically significant PeerEA at age 15 increased from 0% to 37% across MACE MULTIPLICITY scores. This pattern suggests that risk is concentrated in high-multiplicity groups precisely because they contain a higher proportion of individuals who have endured the most potent type-time risk factors.

Our identification of CSA as a predominant risk factor for female substance use is consistent with prior studies and meta-analyses ^2,3^, while the salience of PeerEA in males aligns with a growing body of research on the consequences of bullying ^27,28^. These specific adversities may exert their influence through distinct neurobiological pathways. For instance, we previously reported that PeerEA at age 15 was the strongest risk factor for hyperreactive amygdala response to threatening faces ^11^, and the amygdala is strongly implicated in the neurobiology of SUDs ^29^. Similarly, Pechtel et al. ^30^ proposed that CSA in females produces a specific ecophenotype with deficits in reward anticipation linked to high-risk behaviors, which may help explain its potency as a risk factor.

These results carry significant clinical and public health significance. The absolute increases in risk we observed were striking, often in the range of 20–35%. These risk differences imply that for every three to five exposed adolescents, one additional individual will go on to misuse substances. Consequently, preventing sexual abuse of adolescent girls and peer victimization of adolescent boys could substantially reduce the population burden of substance misuse. From a population perspective, even modest reductions in exposure prevalence could therefore translate into meaningful decreases in the incidence of substance use disorders.

Moreover, identifying adolescents who have experienced these specific adversities may help target interventions to those at greatest risk, whether through early screening, preventive counseling, or an array of trauma-focused therapies^31^.

Several limitations should be noted. First, both childhood maltreatment and substance use histories were based on retrospective self-report, which is subject to recall bias. We sought to mitigate this by using the MACE, a validated instrument with high test–retest reliability^17^, and by verifying exposure and use histories in a substantial subsample through structured interviews. Second, our findings rely on assumptions common to causal inference methods, including exchangeability, no unmeasured confounding, positivity, and consistency. Although these assumptions cannot be fully verified, the integration of TMLE with Super Learner, sensitivity analyses under missing-not-at-random conditions, and calculation of E-values provide reassurance that our results are robust to plausible departures from these assumptions. Third, our sample was restricted to community-dwelling young adults (ages 18–25) from suburban and metropolitan areas, which may limit generalizability to older adults or nationally representative cohorts. However, the narrow age range also confers advantages, as recall of age at exposure is likely more accurate, and this developmental window captures the period of highest risk for substance use onset ^32^. Further, as Dunn and colleagues^7^ have indicated, it is possible to confuse a sensitive period effect with a recency effect; however, the concentration of risk at age 15 rather than ages 17 or 18 argues against recency as the primary determinant. Finally, while our results challenge the cumulative burden model, we do not dismiss the clinical utility of multiplicity as a screening index; rather, we argue that its predictive value is explained by the concentration of the most etiologically salient type–time exposures in high-multiplicity groups. Taken together, these limitations highlight the importance of replication in prospective cohorts, but they do not diminish the strength of our multi-method evidence for mid-adolescence as a sensitive period of vulnerability.

In conclusion, our results, strengthened by modern causal inference methods, highlight mid-adolescence as a developmental window of heightened vulnerability during which exposure to sexual abuse or peer emotional abuse substantially increases risk for illicit substance use. These findings underscore the dual need for prevention strategies targeting these specific forms of maltreatment during this critical period and for increased clinical vigilance in assessing and supporting exposed adolescents.

## Disclosures

Dr. Teicher has received multiple US patents, none of which were used in this study. Dr. Teicher created the Maltreatment and Abuse Chronology of Exposure scale used in this study, but there are no conflicts of interest as he placed the scale into the public domain and made it free to use by all individuals. Dr. Khan reports no financial relationships with commercial interests.

## Data Availability

All data produced in the present study are available upon reasonable request to the authors

## Notes

### Funding Statement

This study was funded by: National Institute on Drug Abuse USA (RO1 DA-017846), National Institute of Mental Health USA (RO1 MH-091391), the ANS Foundation, the Trauma Research Foundation, and Bessel van der Kolk

