## Supplemental materials for "Childhood Maltreatment and Risk for Illicit Substance Use: Evidence for Mid-Adolescence as a Sensitive Exposure Period"

**Appendix**

**Table S1**. Inverse Probability Weighted Cox Proportional Hazard Model statistics

**Figure S1.** Association between the number of different types of maltreatment and adjusted probability of lifetime misuse of various drug classes.

**Figure S2.** Sensitive period analysis of risk factors for illicit substance use in males.

**Figure S3.** Dose-response curves indicating how the risk for use of specific substances varies across levels of perceived financial sufficiency during childhood.

**Figure S3.** Sensitive period analysis of risk factors for illicit substance use in females.

**Table S1**. Inverse Probability Weighted Cox Proportional Hazard Model statistics

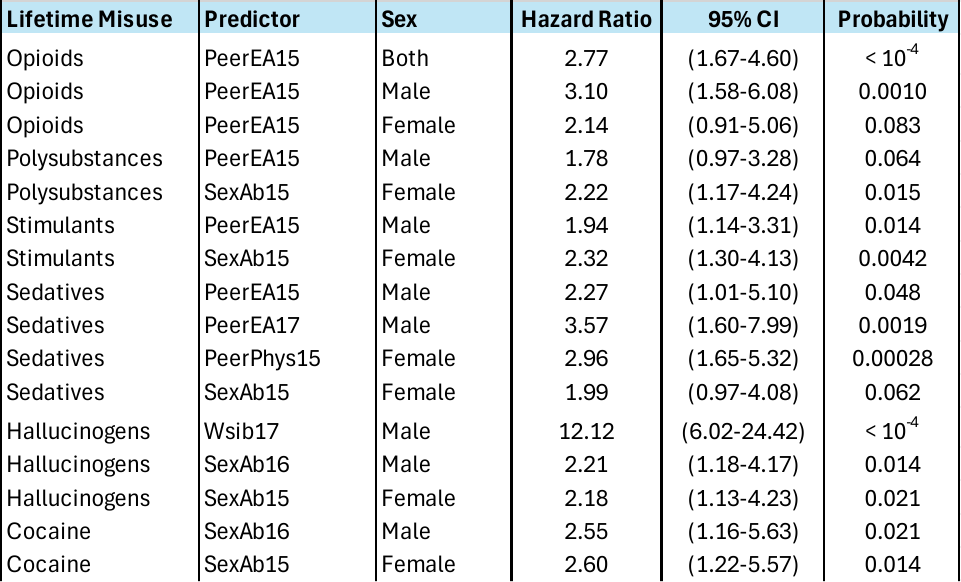

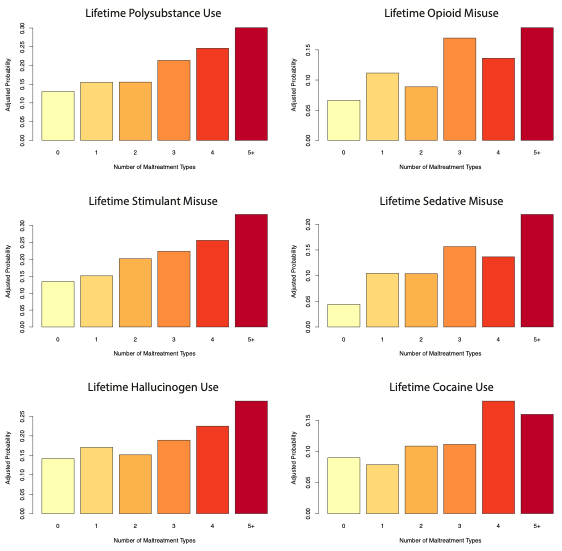

**Figure S1.** Results from logistic regression analysis showing the association between number of different types of maltreatment and the adjusted probability of lifetime misuse of various drug classes. Covariates included age, sex, parental education, and perceived financial sufficiency during childhood.

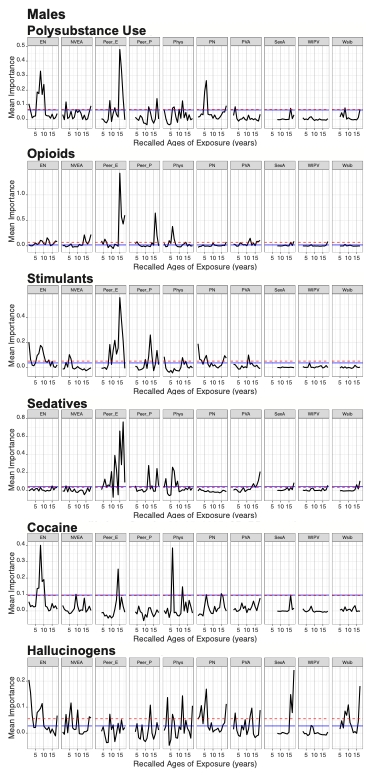

**Figure S2.** Sensitive period analysis of risk factors for illicit substance use in males from random forest regression with conditional inference trees**.** Mean importance is defined as the increase in the mean square error of the fit following the permutation of each variable. The dashed red and solid blue lines indicate risk associated with multiplicity of exposure and severity of exposure, respectively. Abbreviations EN – emotional neglect; NVEA - nonverbal emotional abuse; Peer_E - peer emotional bullying; Peer_P - peer physical bullying; Phys - parental physical abuse; PN - physical neglect; PVA - parental verbal abuse; SexA - sexual abuse; WIPV – witnessing interparental violence; and Wsib – witnessing violence toward siblings. The red dotted line indicates the mean variable importance for MACE Multiplicity (number of different types of maltreatment), and the blue line indicates the mean variable importance of MACE Severity (overall exposure level across childhood).

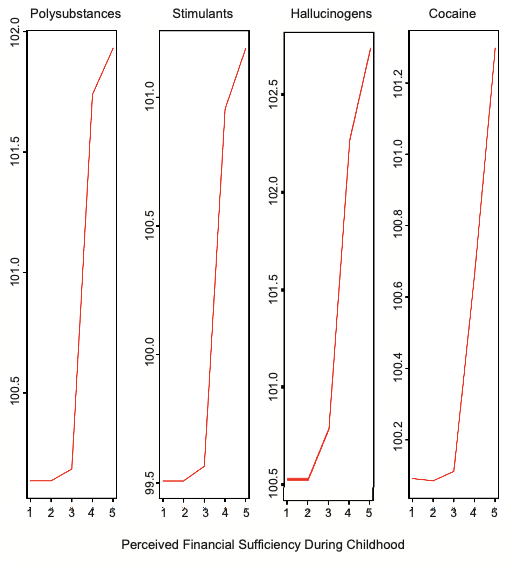

**Figure S3.** Dose-response curves from the saved conditional random forest model. These indicate how risk increases with increasing levels of perceived financial sufficiency, holding all the other risk factors constant at their median level. Perceived financial sufficiency during childhood ranges from 1 – much less than enough money to meet our needs to 5 – much more than enough money to meet our needs.

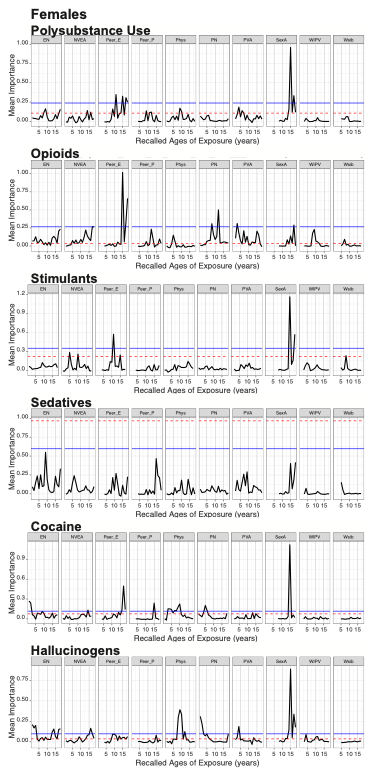

**Figure S4.** Sensitive period analysis of risk factors for illicit substance use in females from random forest regression with conditional inference trees. Mean importance is defined as the increase in the mean square error of the fit following the permutation of each variable. The dashed red and solid blue lines indicate risk associated with multiplicity of exposure and severity of exposure, respectively. Abbreviations EN – emotional neglect; NVEA - nonverbal emotional abuse; Peer_E - peer emotional bullying; Peer_P - peer physical bullying; Phys - parental physical abuse; PN - physical neglect; PVA - parental verbal abuse; SexA - sexual abuse; WIPV - witnessing interparental violence; and Wsib – witnessing violence toward siblings. The red dotted line indicates the mean variable importance for MACE Multiplicity (number of different types of maltreatment), and the blue line indicates the mean variable importance of MACE Severity (overall exposure level across childhood.
